# A Three-Year Retrospective Review of Gynecologic Oncology Referrals to the Specialist Palliative Care Team in a Tertiary Referral Centre: Population, Characteristics, and Outcomes

**DOI:** 10.1101/2024.03.25.24304847

**Authors:** Anthony James Goodings, Mila Pastrak, Sten Kajitani, Elaine Cunningham, Hannah O’Brien, Catherine Weadick, Karie Dennehy

## Abstract

**Background:** The early integration of a specialist palliative care team is demonstrated to have numerous benefits for patients. These extend beyond end-of-life care to include reducing depressive symptoms, improving quality of life, and reducing unnecessary interventions.

**Aims:** This study aims to characterize the patient population referred to the specialist palliative care service with a diagnosis of gynecological cancer. It also assesses referral frequency and response time in order to understand palliative care utilization in an acute hospital setting.

**Methods:** A retrospective chart review and database analysis was performed to extract data on demographics, cancer diagnoses, and referral reasons for patients referred to the specialist palliative care service over three years. The study focuses on identifying patterns in the characteristics of the referred patient population.

**Results:** Analysis of 162 patients revealed a distribution across cancer subtypes: 62% ovarian, 22% endometrial, 12% cervical, and 4% vulvar. A notable finding was that the outcomes for patients with ovarian cancer were more likely to be discharged home with or without community care (61.8%) compared to those with endometrial cancer (41.0%). A rapid response to referrals was observed, with 70% reviewed within three days and 98% within a week. This highlights the service’s efficiency and the demographic and diagnostic profile of the patient served.

**Conclusions:** This study gives insight into the demographic and diagnostic profiles of gynecological cancer patients referred for palliative care, alongside demonstrating rapid response to such referrals. Despite the rapid assessment times, the research importantly identifies differences in outcomes among different cancer subtypes, with a particular emphasis on the variance in discharge destinations. These findings reflect both patient preference and medical need, demonstrating the role of tailoring palliative care approaches to meet the individual needs and desires of this diverse patient population.

## Introduction

The treatment of gynecological cancer as well as many other types of cancers has drastically improved over the past several decades, indicating that more people are cured and have longer lifespans with the disease (1). As a result, this has led to a need to adapt and reconsider the role of palliative care (2) and has been universally reflected in the change of the WHO’s definition of palliative care, 2002: “palliative care improves the quality of life of patients and that of their families who are facing challenges associated with life-threatening illness, whether physical, psychological, social or spiritual” (3). Further, the WHO considers that palliative care should be incorporated “from early in the disease course, can be delivered alongside potentially curative treatment, and continues to include end-of-life or terminal care” (4). Palliative care attempts to alleviate the suffering of patients is through early identification and treatment of new symptoms, while also managing refractory symptoms. However, palliative care needs to be embraced by all medical providers, including oncologists and other team members involved in the care of the patient, in addition to the palliative care physicians and nurses.

The shift towards an integrated care model, including both the oncology team and a specialist palliative care team has been associated with several benefits to patients (5). In addition to end-of-life concerns, palliative care integration can address long-term problems related to disease and treatment of gynecological cancers. Specifically, early integration of palliative care has been found to improve quality of life (6), reduce depressive symptoms (5), reduce unnecessary invasive treatment in the last six weeks of life, and was determined to be an independent prognostic factor for overall survival (5).

The American Society of Clinical Oncology endorses the early use of palliative care within clinical oncology as it has shown that palliative care intervention improves quality of life, is cost effective, and does not adversely affect patient outcomes (7), yet palliative care for gynecological cancer is still underused (8,9). Its integration is associated with various social benefits such as increased likelihood of a structured end-of-life discussion (9) and markers of increased quality of life, such as more home deaths and less non-beneficial chemotherapy use (5,9). Gynecological cancer in particular carries additional complications including fertility issues and sexual side effects which require specific management (10). Thus, the need for social and psychological support is beneficial in treating most patients with gynecological cancer (11). A palliative care team as a part of an integrated care model is well suited to deliver this.

A lack of awareness and comprehensive understanding of the services provided by palliative medicine teams is one of the primary barriers to referral (12). While oncologists integrate components of palliative care, such as addressing the symptomatic involvement including nausea, vomiting, pain, anorexia, diarrhea, electrolyte imbalances, etc., patients should have palliative involvement to assist in appropriately managing complex cases. Further, the timing of referrals to palliative care continues to be an obstacle in providing appropriate support to patients (12). Evidence shows that patients with advanced gynecological cancer are referred late in their diagnosis and treatment course (13). The current approach to management of gynecological cancer often incorporates palliative care on a case-by-case basis rather than through a standardized framework adapted to the purpose. As such, patients can end up having suboptimal clinical outcomes relating to their quality of life and symptom control. As a result, the underutilization and poor implementation of palliative care often leads to difficulties in avoiding aggressive interventions in the later stages of life.

Building a strong rapport between the patient with gynecological cancer, their oncologist, and the other members of the multidisciplinary team can provide a foundation for understanding the need for a palliative perspective during the disease course, despite the preconceptions surrounding it (9). Because of the known and extensive benefits of early involvement of a specialist palliative care team as a part of an integrated care model, an understanding of the frequency and reasons for palliative care referral can help further encourage the use of the specialist palliative care service.

### Aims and Objectives

The aims of this study are to understand the patient profile, indication for referral, and outcomes of patients with gynecological cancer referred to the palliative care service.

1. **Characterizing the Patient Population**: The goal is to delineate the demographic and diagnostic profiles of patients with gynecological cancers who are referred to the palliative care service, aiming to identify patterns that may inform future care strategies.
2. **Analyzing Referral Outcomes**: This objective seeks to examine the outcomes of patients who are referred to the palliative care service, and determine outcomes such as: discharged home with or without community care, discharged to hospice care, or death in hospital.

The objective of this study is to assess current practice and identify areas where better integration of palliative care services can lead to improved outcomes for patients with gynecological cancer. By reviewing current practices, we aim to discover gaps and pinpoint opportunities for enhancement, thereby improving the quality of care and support for these patients. The ultimate goal is to enhance patient outcomes and quality of life and potentially extend survival times.

## Methods

The study aims to retrospectively analyze referrals for patients with gynecological cancer to the inpatient consult palliative medicine service at an acute hospital (Cork University Hospital) over a three-year period. The following is an overview of the methods that was employed to achieve this objective:

1. Data Collection: A retrospective chart review and database data extraction was conducted to gather data on patient demographics, diagnosis, time to first review by the service, duration of ongoing review by palliative care team in hospital, and patient outcomes (transfer to hospice, discharge from service, community care, or death).
2. Data Anonymization: The collected data was pseudonymized to protect patient confidentiality while still being able to determine re-referrals and duplicate database entries.
3. Data Storage: The data was stored on a password-protected computer using SPSS and UCC OneDrive For Business.
4. Data Analysis: Statistical analysis involved descriptive statistical computation using SPSS version 28.

Ethical approval was obtained from the Clinical Research Ethics Committee of the Cork Teaching Hospitals.

### Population

The study encompassed all patients with gynecological cancer referred to the specialist palliative care service from January 2020 to January 2023, a three-year period.

#### Inclusion criteria

1. Adult patients with a confirmed diagnosis of a gynecological cancer.
2. Patients who were referred to the hospital palliative care service between January 2020 to January 2023.

#### Exclusion criteria

1. Patients under the age of 18 years.
2. Patients with non-gynecological cancer diagnoses.
3. Patients who were not referred to the palliative care service during the specified time period.

### Recruitment

The specialist palliative care team maintains a record of all patients reviewed by and referred to the service, which was cross-referenced through the directorate data manager of an online database maintained by Cork University Hospital Oncology. All referrals to the palliative care service were included or excluded based on the criteria above. In case of any discrepancy between the palliative care service and data manager patient lists, a review of the relevant patient chart was conducted to determine eligibility.

### Research Tools

The primary tools for this study included a database of patient records for all inpatients referred to the specialist palliative care team at Cork University Hospital. The database was accessed to gather data on patients diagnosed with:

- Any type of cancer originating from or strongly associated with the female reproductive system, including cervical, ovarian, endometrial (uterine), vaginal, vulvar, gestational trophoblastic disease, fallopian tube, primary peritoneal, and Müllerian carcinomas.

Data collection encompassed a time frame of January 2020 to January 2023.

### Variables

Data collected included: Medical record number (MRN) – pseudonymized, age, diagnosis, time to first review by the service, duration of ongoing review by the specialist palliative care team in hospital, and patient outcome.

### Data Analysis

1. Statistical Analysis: SPSS software version 28 was used for descriptive statistical analysis, categorizing patients by cancer subtype and their indication for referral. This formed distinct patient cohorts.
2. Referral Proportion: The number of gynecological malignancy referrals were quantified as a proportion of total referrals to the palliative care service over a three-year period.
3. Discharge Outcome: For patients who were discharged from the service, follow-up plans in the data collected were included, such as whether community palliative care services were requested on discharge.

A detailed review of individual patient charts were conducted to understand instances where the outcome was not clearly documented on the database.

## Results

Over the three-year period examined, a total of 162 individuals with a gynecological cancer diagnosis accessed the inpatient specialist palliative care service at Cork University Hospital. This population was described using several metrics, including diagnosis, outcome, duration of care, time to being seen, and age.

**Table 1:** Subtypes of Gynecological Cancers and their Respective Referrals.

| Subtype | Proportion of Referrals | Number |
| --- | --- | --- |
| Ovarian | 59.0% | 95 |
| Endometrial | 24.8% | 40 |
| Cervical | 11.2% | 18 |
| Vulvar | 5.0% | 8 |

**Table 2.** Outcomes Respective to Subtypes of Gynecological Cancers. D/C: discharged CC: community care SPCU: Specialist Palliative Care Unit w/o: with out

| Outcome | Ovarian | Endometrial | Cervical | Vulvar |
| --- | --- | --- | --- | --- |
| D/C Home + CC | 50.6% (n=45) | 41.0% (n=16) | 55.6% (n=10) | 85.7% (n=6) |
| SPCU | 28.1% (n=25) | 46.2% (n=18) | 38.9% (n=7) | 14.3% (n=1) |
| Death (Hospital) | 10.1% (n=9) | 12.8% (n=5) | 5.6% (n=1) | 0 |
| D/C Home w/o CC | 11.2% (n=10) | 0 | 0 | 0 |

The mean duration that patients were followed by the inpatient team was 7 days, with a range of 1 to 55 days (SD ± 8.7 days). The service saw 70% of referred patients within 3 days and 98% within one week.

The mean age of patients seen by the service was 65.8 years with a standard deviation of 13.8 years.

When assessing outcomes, it was noted that patients with ovarian cancer were more likely to be discharged home with community care (50.6%) or admitted to a specialist palliative care unit outside of the acute hospital (28.1%), compared to patients with endometrial cancer (41% and 46.2%, respectively). Patients with cervical or vulvar cancer had varying outcomes, with a higher proportion being discharged home with community care (55.6% and 85.7%, respectively), although the sample size was small in these two groups (18 and 7, respectively).

**Figure 1:**
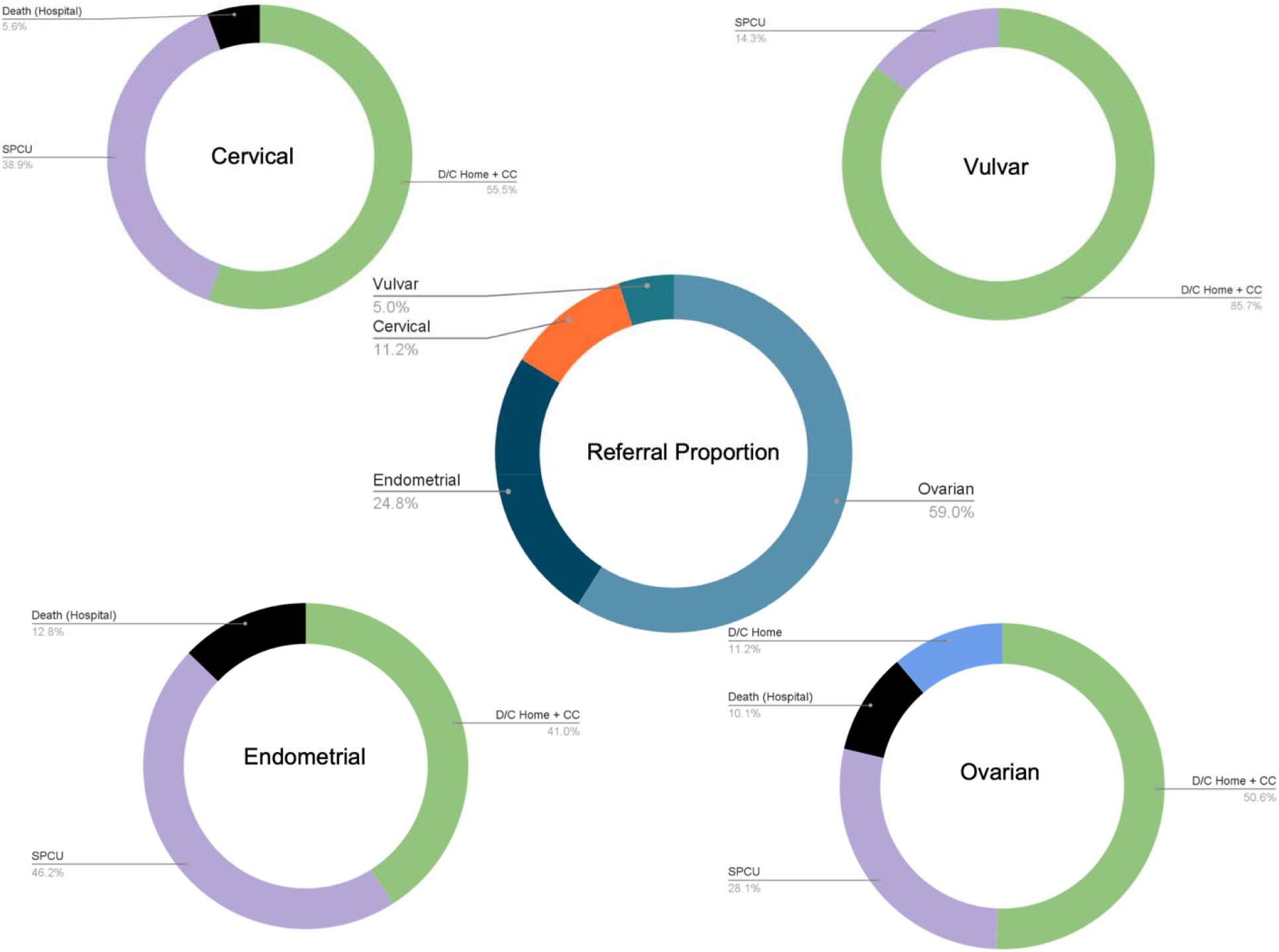
Referral Proportion and Outcomes

## Discussion

This retrospective study has aided in the attainment of a better understanding of the population of patients with gynecological cancer being referred to the specialist palliative care service at Cork University Hospital. By exploring these specific cases, we were able to identify potential factors contributing to variations in patient outcomes when engaging with the palliative care service. The referral proportion demonstrated that the majority of patients referred to the service had ovarian cancer, with endometrial being the next most common. Over a three-year period, the service encountered patients with cervical or vulvar cancer less frequently. The mean duration of care provided by the inpatient team was relatively short (7 days), with a wide range (1 to 55 day) indicating variability in the length of time patients received inpatient palliative care services. Studies on the subject suggest that prompt access to care is essential (2,12). This was well achieved in the cohort of patients reviewed in this study with 98% of patients referred being seen within a week.

The findings regarding outcomes for patients with gynecological cancer highlight several important considerations for palliative care delivery and service optimization. Patients with cervical or vulvar cancer had varying outcomes, with a higher proportion being discharged home with community care although the sample size was low. The data shows that patients with ovarian cancer are more likely to be discharged home with or without community care compared to patients with endometrial cancer. This discrepancy likely reflects disease trajectory and patient desires, highlighting the importance of an individualized approach and integration of a multidisciplinary team to support the patient in the setting that is most comfortable to them.

Patients discharged without community follow-up may result in challenges in ensuring comprehensive symptom management and supportive care post-discharge. Patients discharged home are generally offered community care, including pain management, psychosocial support, and rehabilitation, which are crucial components of palliative care delivered by a multidisciplinary team. Should the patient reject the referral to community care, their oncologist generally re-referred them to the service at a later time. This is done to prevent inadequate post-discharge support as it may lead to higher rates of readmission and poorer quality of life in this patient population (14)

Hospitals can implement several strategies to optimize the management of gynecological cancer patients and improve their post-discharge outcomes. Firstly, hospitals should prioritize comprehensive pre-discharge assessments to identify patients’ specific needs and desires in order to develop individualized care plans. This demands a comprehensive assessment of individual patient profiles, encompassing considerations such as post-surgical recovery status, analgesia efficacy, and risk of complications. This approach requires conducting in-depth symptom evaluations, delivering patient education regarding self-care techniques, and facilitating the integration of community-based support resources, including home healthcare services and rehabilitation programs.

Multidisciplinary teams can help improve patient outcomes through collaboration between palliative medicine physicians, oncologists, surgeons, nurses, social workers, and other allied health professionals (15). By providing comprehensive education to patients and caregivers on postoperative care instructions and signs of potential complications, hospitals can empower patients to actively participate in their recovery process (16).

Finally, it may be of benefit to consider the use of telemedicine in gynecologic cancer care as it can improve accessibility, reduce costs, and ensure continuous support for patients - particularly those in remote areas (17) who would prefer to receive home care. Additionally, research has highlighted that telemedicine can improve access to palliative care services, thereby reducing subsequent hospitalizations among gynecological cancer patients (17).

This study has focused on patients with gynecologic cancer who were referred to the specialist palliative care team, based solely on available recorded data. As such, it does not account for those not referred to the team within the same period. The retrospective nature of this analysis means that findings are limited to the scope of existing documentation. While these considerations are important, they do not significantly detract from the overall insights gained from the study.

## Conclusion

In conclusion, this retrospective study at an acute tertiary hospital over a three-year period provides valuable insights into the specialist palliative care needs of patients with gynecological cancers, predominantly ovarian and endometrial cancers. It underscores the variability in care duration and the efficacy of prompt palliative care interventions, with a significant majority of patients referred being assessed within a week of referral. The outcomes highlight the importance of a tailored, multidisciplinary approach to palliative care that considers individual patient profiles, including postoperative recovery, pain management, and potential complications. The findings advocate for comprehensive pre-discharge planning, community-based support integration, and the potential benefits of telemedicine to enhance palliative care accessibility and patient outcomes. This study reinforces the need for individualized care plans and the value of a multidisciplinary team in optimizing the quality of life for patients with gynecological cancers. Further research could include exploring the patient experience with the specialist palliative care team.

## Data Availability

All data produced in the present study are available upon reasonable request to the corresponding author.

## Acknowledgements

This project owes its success to the contributions of several individuals and supporting institutions, whose expertise and guidance have been invaluable. We extend our heartfelt thanks to Cork University Hospital and University College Cork for providing access to the facilities and resources necessary for conducting this study. Special recognition is given to Dr. Karie Dennehy, Consultant in Palliative Care Medicine, for her supervision and invaluable insights throughout the project. We are also grateful to Prof. Seamus O’Reilly, Oncology professor, for offering critical feedback on the study design. Their collective support has been instrumental in advancing our understanding of palliative care utilization for patients with gynecological malignancies.

This project did not receive any internal or external funding. The research and all associated activities were carried out with the support of existing resources provided by the affiliated institutions and the voluntary contributions of the team members involved.

